# Intensity of COVID-19 in care homes following Hospital Discharge in the early stages of the UK epidemic

**DOI:** 10.1101/2021.03.18.21253443

**Authors:** Joe Hollinghurst, Laura North, Chris Emmerson, Ashley Akbari, Fatemeh Torabi, Ronan A Lyons, Alan G Hawkes, Ed Bennett, Mike B Gravenor, Richard Fry

**Affiliations:** Population Data Science and Health Data Research UK, Swansea University; Public Health Wales; Hawkes Centre for Empirical Finance, Swansea University; Computer Science, Swansea Univeristy; Swansea University Medical School, Swansea University

**Keywords:** Care homes, Hospital discharge, COVID-19, Linked Data, Multi-level model, Hawkes Process

## Abstract

**Background:** A defining feature of the COVID-19 pandemic in many countries was the tragic extent to which care home residents were affected, and the difficulty preventing introduction and subsequent spread of infection. Management of risk in care homes requires good evidence on the most important transmission pathways. One hypothesised route at the start of the pandemic, prior to widespread testing, was transfer of patients from hospitals, which were experiencing high levels of nosocomial events.

**Methods:** We tested the hypothesis that hospital discharge events increased the intensity of care home cases using a national individually linked health record cohort in Wales, UK. We monitored 186,772 hospital discharge events over the period March to July 2020, tracking individuals to 923 care homes and recording the daily case rate in the homes populated by 15,772 residents. We estimated the risk of an increase in cases rates following exposure to a hospital discharge using multi-level hierarchical logistic regression, and a novel stochastic Hawkes process outbreak model.

**Findings:** In regression analysis, after adjusting for care home size, we found no significant association between hospital discharge and subsequent increases in care home case numbers (odds ratio: 0.99, 95% CI 0.82, 1.90). Risk factors for increased cases included care home size, care home resident density, and provision of nursing care. Using our outbreak model, we found a significant effect of hospital discharge on the subsequent intensity of cases. However, the effect was small, and considerably less than the effect of care home size, suggesting the highest risk of introduction came from interaction with the community. We estimated approximately 1.8% of hospital discharged patients may have been infected.

**Interpretation:** There is growing evidence in the UK that the risk of transfer of COVID-19 from the high-risk hospital setting to the high-risk care home setting during the early stages of the pandemic was relatively small. Although access to testing was limited to initial symptomatic cases in each care home at this time, our results suggest that reduced numbers of discharges, selection of patients, and action taken within care homes following transfer all may have contributed to mitigation. The precise key transmission routes from the community remain to be quantified.

## Introduction

Care homes are a cornerstone of adult social care which, by definition, group clinically vulnerable people together. Residents may live in close proximity, access shared space, and may be frail and experience a variety of underlying health conditions, making them more susceptible to outbreaks of infectious disease than those who live independently (1). COVID-19 is described by Lithander et al (2020), as ‘a dynamic, specific and real threat to the health and well-being of older people’. The impact of COVID-19 on this sub-population was a defining feature of the pandemic, reported internationally by care providers, in the media, and through a growing body of research (2,3). Most care homes formed protective bubbles whereby visitors were restricted to essential staff and the admission of new residents. Yet there were still many outbreaks and deaths from COVID-19, and there remains limited evidence on modifiable risk factors for increases in COVID-19 infections in care homes. Regularly, attention was drawn to links between hospital acquired infection and discharge into the care home (4).

Care homes provide accommodation and care for those needing substantial help with personal care, but more than that, they are people’s homes (1,5). In 2016, there were 11,300 care homes in the UK, with a total of 410,000 residents (6). Care home markets vary across the 22 local government authorities in Wales in the supply, ownership, and size of care homes (7). Following a wide reaching inquiry into quality of life and care in care homes, the Older Person’s Commissioner for Wales concluded that ‘too many older people living in care homes have an unacceptable quality of life’ (8). The Commissioner’s expectations for change were far ranging and included greater investment in the care home sector, staff development, recalibrating a human rights focus, quality reporting and provision of a range of health services (8).

Multiple interconnected challenges face the care home sector in the prevention and management of outbreaks of COVID-19 (5). In the literature, these challenges are reported to include staff shortages (5,9), insufficient or lack of timely COVID-19 testing and poor access to personal protective equipment. Related clinical challenges include older adults with COVID-19 being asymptomatic, or not displaying expected symptoms (5,9–11). Once there is an outbreak, the disease can spread quickly within a care home setting, and be difficult to contain (10,12,13). A further challenge is in managing the impact of practices to shield care home residents and isolate those who are infected. These practices can result in social isolation from families, friends and communities, with negative impacts on health and wellbeing (5,11). Set against these challenges is the caring, innovative, and resilient response of care home staff and residents in managing the situations they face (14).

This confluence of events in the context of the pandemic, and impacts for residents, their families and care home staff, has been framed as a human rights issue (15). In the UK it is argued that underinvestment in the care home sector and a poor interface with the health sector led to ill-informed policies. The rapid hospital discharge policies in the early period of the lockdown have been presented as examples of this (13,16–18). However, this was a period of considerable uncertainty, with very limited testing, and rapidly increasing transmission in all communities. A quantitative estimate of the effect of this route of transmission is therefore difficult to obtain.

The use of existing anonymised routinely collected longitudinal data can help to provide rapid access to large-scale data for studies and provide robust evidence for commissioning decisions and policy (19). In this study, we utilise the Secure Anonymised Information Linkage (SAIL) Databank (20–22) to investigate increases in confirmed COVID-19 cases in care homes in Wales, in periods of time following a recorded exposure from a patients discharged from hospital.

Understanding the pathways in which a virus has entered a community is key to preventing the spread of disease, particularly when the community vulnerable. Logically, in a controlled environment like a care home there are four routes of ingress for the virus: hospital discharge, staff, visitors, and community admissions. This study addresses the first of these routes of ingress by assessing the impact of hospital discharge of COVID-19 patients into care homes on subsequent COVID-19 cases and whether any of the care home characteristics increased the risk.

## Methods

### Data sources

We performed a national, longitudinal, retrospective cohort study using the SAIL Databank, a person level anonymised privacy protecting data linkage platform for all >3 million people who live in Wales, UK (20–22). SAIL contains anonymised administrative and healthcare records. Anonymisation is performed by a trusted third party, the National Health Service (NHS) Wales Informatics Service (NWIS). A unique individual anonymised person identifier (Anonymous Linking Field, ALF) links to a unique address anonymised identifier (Residential Anonymous Linking Field, RALF, (23,24). Individual linking fields, nested within residential codes, are contained in the anonymised version of the Welsh Demographic Service Dataset (WDSD), replacing the identifiable names and addresses of people registered with a free-to-use General Practitioner service.

Our care homes anonymised linking field (CHALF) was created using the Care Inspectorate Wales (CIW), the national regulator of social care services in Wales, list from 2020, and assigning a Unique Property Reference Number (UPRN, (25)). The UPRN was double encrypted into a project-level RALF, and uploaded into SAIL to create a deterministic match to the WDSD based on patient supplied addresses. Additional characteristics of care homes were supplemented using Geographical Information Systems data. We determined if someone was a care home resident by linking their anonymised address information to the residences indexed as a care home in the WDSD. This enabled us to link COVID-19 testing and hospital discharge data at the individual level, and within individual care homes. Residents and care homes were anonymised prior to any analysis.

Our dataset consisted of 928 care homes with at least one current resident, successfully linked to anonymised patient data on 15,772 residents. Daily observations on case numbers of hospital transfers were made from 1^st^ January 2020 to 31^st^ July 2020, a dataset of 186,772 observations. We were unable to include residents who were temporarily discharged from hospital to a care home different to their normal place of residence. We carried out two sets of analysis. (1) A multi-level logistic regression estimate of the association between exposure to hospital discharge and the risk of cases subsequently increasing in the care home. (2) A novel stochastic outbreak model, to estimate the change in intensity of cases in a care home following the date of exposure, and taking into account the nature of spread within the home.

### Multi-level modelling

We defined the outcome in a binary logistic regression model as the increase in COVID-19 positive cases, *c*, in a care home, defined daily by the difference in the sum of cases across a two-week moving window:

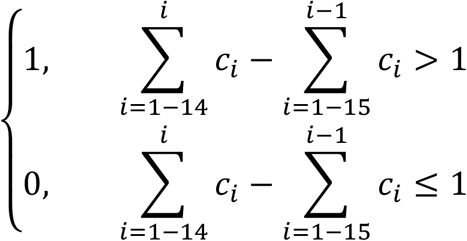

14 days is the 99^th^ percentile of an individual’s symptomatic period (26) and is the recommended quarantine period in the UK. As a marker of exposure to hospital discharge, we included a time-dependent covariate, defined as “yes” if a resident of the care home was discharged from hospital in the previous 14-days. Fixed effects of type of nursing provision, mental health provision, learning disability provision, home capacity (defined by number of available beds recorded with CIW), and resident density were included. To account for clustering, observations were nested within month, home then local authority. We varied the periods of observation as a sensitivity analysis.

### Hawkes Process Model

For an infectious disease in a small, closed population, there is clearly strong non-independence between observations on cases over time. We therefore developed a novel stochastic model to attempt to separate out the effects of introduction of a case from the subsequent spread within the home. An assignment of potential introduction via the hospital discharge route can then be estimated separately from both the within-home spread, and a ‘background’ introduction rate from other outside sources (community/staff/visitors).

A random point process model for the discrete event of a confirmed case can be expanded upon using the Hawkes process (Hawkes, 1971). The feature of this model is a self-exciting term representing a situation in which the probability of a subsequent cases is increased, or ‘excited’ by an existing case. The period of excitation represents an outbreak of, potentially, many cases within a care home. We proposed an outbreak model with two self-excitation terms: one representing the effect of a known case happening in the home (a considerable risk to future spread), and the other one representing the effect of a known hospital discharge. Since the hospital discharge may or may not carry infection, the relative magnitude of these terms informs an estimate of the risk from hospital discharge. In the extreme case, zero hospital excitation would represent a situation where no hospital discharges carried infection risk (null hypothesis), while equal excitation coefficients represent the situation where every hospital discharge was infected.

The *intensity* of cases is the rate at which cases are expected to occur. We defined the *intensity function* of cases at care home *i* on day *t* as:

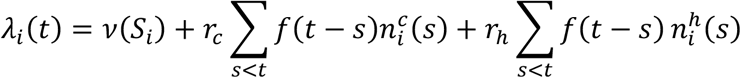

where *v*(*S*_*i*_) is the baseline intensity of a care home of size *S*_*i*_, representing the risk of a case being introduced to the care home via normal activities (staff/community/visitors), *r*_*c*_ and *r*_*h*_ are the self-excitation and excitation by hospital parameters, representing the increased risk of an outbreak following the introduction of a case or hospital transfer respectively, *f*(*t*) is the COVID-19 serial interval distribution, assumed equal to the gamma probability density function with mean 6.5 and coefficient of variation 0.62, and 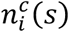 and 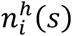 are the numbers of cases in and hospital discharges to the care home *i* on day *s*. The care home size *S*_*i*_ is grouped into quartiles such that there are four possible values for *v*(*S*_*i*_).

We assume *n*_*i*_(*t*) has a Poisson distribution with mean *λ*_*i*_(*t*), hence the model can be fitted from the observed time series of hospital discharge and case time series. We used maximum likelihood and MCMC using Numpy (Harris et al., 2020, Bennet, 2020) for three models: with *r*_*c*_ and *r*_*h*_ both fixed to zero, with only *r*_*h*_ fixed to zero, and with no parameters fixed. Details of the MCMC fit are given in supplementary material along with an illustration of the effect of the self-exciting process on the intensity of cases (Figure S1).

## Results

Care home characteristics are summarised in table S1 (supplementary information). There was no clear temporal association between cases recorded in care homes and numbers of hospital discharges over the period April-June 2020 (figure S2, supplementary information). In our two analytical approaches, we found estimates of no effect, and a small effect of hospital discharge on subsequent COVID-19 case rates in the care homes.

### Multi-level Modelling

In unadjusted univariable analyses, (Table S2, supplementary material), the marker of hospital discharge was associated with an increased risk of a rise in cases in the care home (odds ratio (OR) 1.2, 95% CI 1.0, 1.5). By far the biggest risk factor was care home size (OR 34.6 for the largest quintile in comparison to the smallest). As expected, there was a correlation between hospital discharge and care home size, and in the adjusted multivariable models (Table 1), there was no significant effect of hospital discharge (OR 0.99, 95% CI 0.82, 1.90). Care home size remained the most important factor in mutually adjusted analyses. In general, we found provision of learning difficulty to reduce the risk, the provision of nursing increased risk, while the provision for mental health had no effect. Risk was also increased as the density of residents increased. The random effects terms indicated that an increase in COVID-19 cases varied significantly by month (largest residual) and by care home, but not by local authority.

**Table 1.**
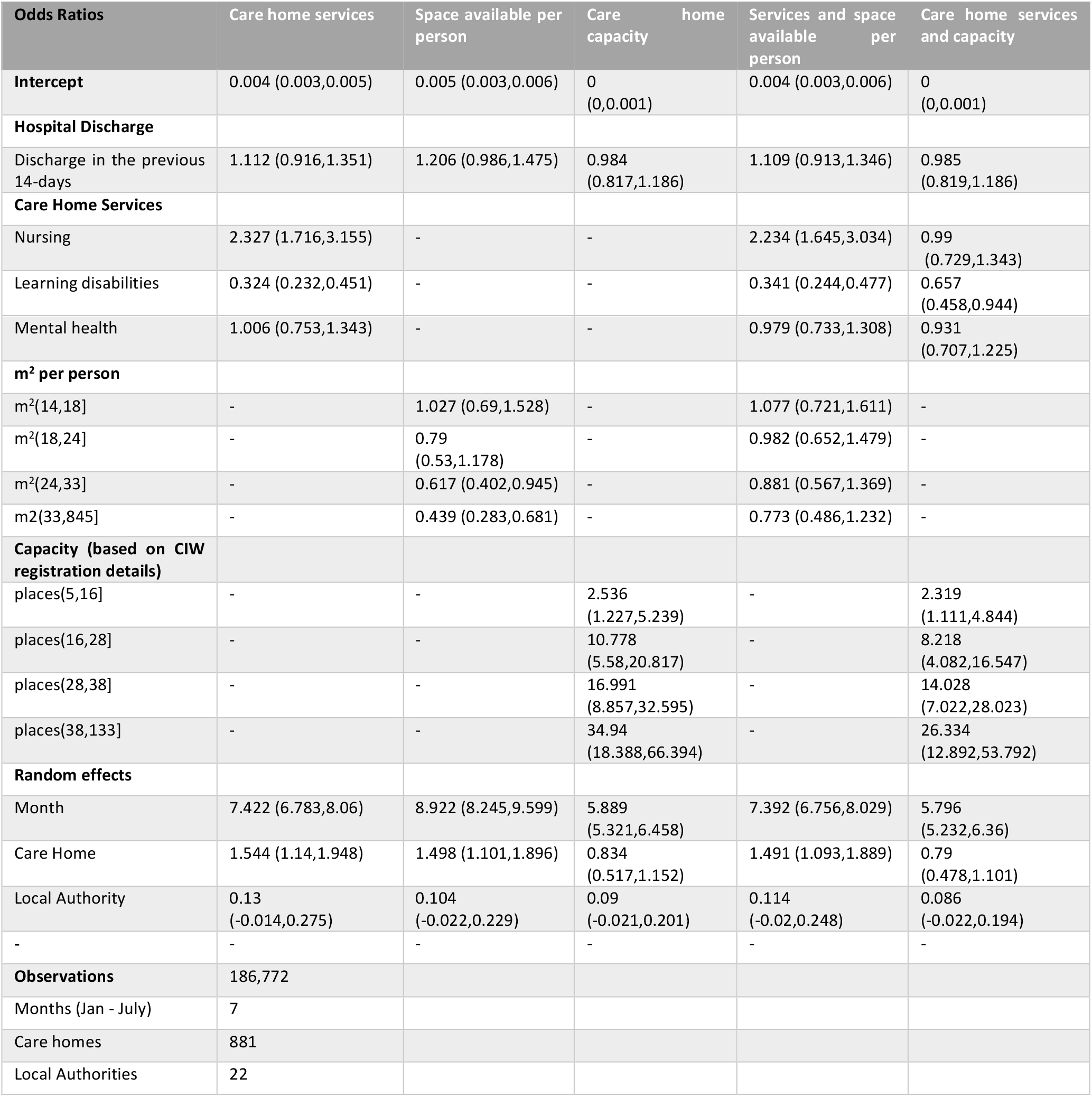
Odds Ratios for the multivariate multilevel logistic regression models, with the dependent variable being an increase in COVID-19 positive cases in a care home.

### Hawkes Process Model

The inclusion of the case self-excitation term led to a significant improvement in the fit of the model over the care home size only model (p < 0.001), with the magnitude of the self-excitation effect considerably larger than the care home size effect. We found a further significant improvement in the likelihood of the model with hospital discharge, compared to the model with case excitation only (p < 0.001). The magnitude of change in intensity associated with hospital discharge was comparable to the baseline intensity in a Q2 care home, and considerably smaller than the baseline effects of Q3 or Q4 homes. Furthermore, the case self-excitation coefficient was much larger. We estimated the ratio *r*_*h*_ / *r*_*c*_ = 0.018, suggesting one hospital discharge had the same effect on intensity as 0.018 cases. An alternative interpretation would be that 1.8% of hospital discharges may have been infected. Estimates of the parameters from the full model are shown in Table 2. Details of the MCMC fit are given in supplementary material (Figure S4).

**Table 2.**
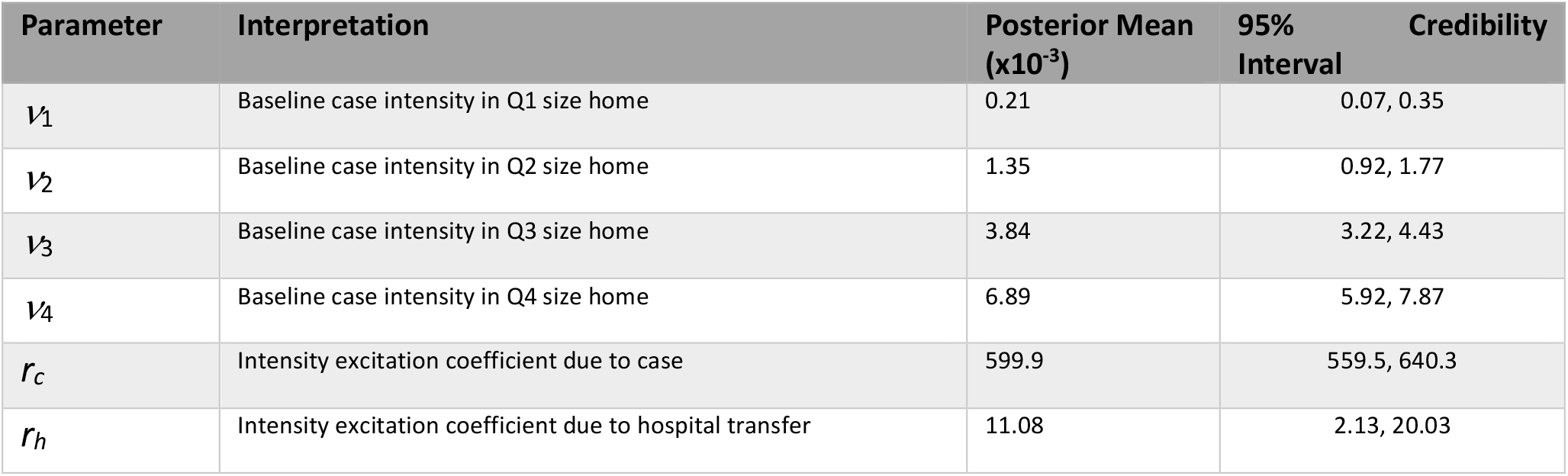
MCMC estimates of parameters for the full Hawkes process model.

## Discussion

We used two modelling approaches, in combination with a national individually linked hospital and care home event cohort, to explore the role of hospital discharge into care homes on subsequent COVID-19 case rates. The study focused on the period, during the first wave in the UK, during which this was most likely to have been a factor, before widespread testing was available. The results of both approaches suggest only a minor role for this transmission pathway, which has attracted much comment and speculation. In our multi-level regression, we found no significant effect of recent hospital discharge on the probability that case levels would subsequently increase. Care home size, and provision type (specifically nursing provision) were identified as risk factors. We were also able to characterise care homes by number of residents per m^2^ based on building footprint, and show that high person density was a risk factor likely representing the increased opportunity for spread of directly transmitted viruses within a closed population.

Our hierarchical regression models attempted to control for effects of clustering, however by its nature there is a very strong likelihood of a direct link between cases of infectious disease located closely in time and space. We aimed to account for this more explicitly by modelling the process of introduction into the care home, and subsequent spread by developing a simple stochastic epidemic model whereby the occurrence of a case is self-exciting, and leads to an increased probability of further cases. Fitting the model with case self-excitation only, resulted in a considerable improvement over the size-only model. This was a ‘sense-check’ result, as it was essentially a test of the hypothesis that SARS-COV-2 was highly infectious within a care home: if a case occurred, there was a highly significant increase in the risk of more cases over the following days. The magnitude of the effect was considerably larger than the care home size effect. This also highlights the importance of including the infectious dynamics in the analysis. Without it, the effect of care home size, or any other variable, may be over-estimated.

When we included the impact of hospital discharge, we did indeed find a significant effect. If a care home was exposed to a discharge event, there was an estimated increase in the intensity of cases. However, the effect was relatively small. In comparison with the introduction of a known case, we estimated an approximately 50-fold lower impact. One way of interpreting the coefficients would be that an estimated 1.8% of patients discharged from hospital into a care home may have been infected. The hospital effect was considerably less than the effect of care home size. We estimate that the change in risk posed by one hospital discharge event *every week* was equivalent to the change in risk comparing a Q2 sized care home to a Q1 sized home, and much less than the difference between larger home sizes (Figure S4, supplementary material).

Data linkage is a powerful tool for building comprehensive cohorts. However, whilst SAIL allows us to model a person’s care pathway and outbreaks at an individual care homes, it is reliant on timely updates of a person’s General Practice (GP) record. During wave one of the COVID-19 pandemic, when social-care and health policy was being revised on a weekly basis, we know temporary discharge events to care homes took place, but we are unable to include these cases in our analysis. We were unable to link all care home residents to care homes in Wales due to mismatches in GP recorded addresses and officially registered care home address (∼10% of care homes). This may have led to some missed discharge events, but at the population level of this study this is unlikely to have drastically influenced the resulting analysis.

## Conclusions

Our results agree with other studies (2,3), but are the first to use individual case events as the outcome. Our model is generalisable to other discrete exposure events, and subsequent disease spread, and can also be developed to include other risk variables. There has been a natural tendency to link hospitals and care homes as a transmission route for COVID-19. During the first wave, there was pressure to maximise hospital bed availability as the peak of the epidemic was approaching, and there was a lack of readily available rapid testing. However, when taken as a whole, the evidence around discharge from hospitals into care homes suggests that most care home outbreaks were related to community infection either from visitors, visiting professionals or staff. Given the high prevalence in UK hospitals at the time, this suggests that successful mitigation was put in place through the (pre-testing) decision making regarding transfers, the substantial decline in discharge rate that was introduced prior to the first wave epidemic peak, and management of patients once transferred into the care home.

## Data Availability

The data used in this study are available in the SAIL Databank at Swansea University, Swansea, UK. All proposals to use SAIL data are subject to review by an independent Information Governance Review Panel (IGRP). Before any data can be accessed, approval must be given by the IGRP. The IGRP gives careful consideration to each project to ensure proper and appropriate use of SAIL data. When access has been approved, it is gained through a privacy-protecting safe haven and remote access system referred to as the SAIL Gateway. SAIL has established an application process to be followed by anyone who would like to access data via SAIL https://www.saildatabank.com/application-process. Findings will be presented at conferences, published in peer-reviewed journals and to the funders and government COVID-19 advisory bodies as appropriate. Strengthening the Reporting of Observational Studies in Epidemiology (STROBE) and Reporting of studies Conducted using Observational Routinely-collected Data (RECORD) (via the COVID-19 extension) checklists will guide our study findings reporting.

http://www.saildatabank.com

## List of abbreviations

ALF: Anonymous Linking Field
CHALF: Care Homes Anonymised Linking Field
CIW: Care Inspectorate Wales (CIW)
NHS: National Health Service
NWIS: NHS Wales Informatics Service
OR: Odds Ratio
RALF: Residential Anonymous Linking Field
SAIL: Secure Anonymised Information Linkage
UPRN: Unique Property Reference Number
WDSD: Welsh Demographic Service Dataset

## Author’s Contributions

RF, JH, CE, MBG, RAL conceived the study design, performed the data creation and linkage, analyses and drafted the paper. AH, EB, MBG developed the Hawkes modelling analysis with input from JH, RF and LN. All authors contributed to data curation and linkage. All authors gave final approval of the version to be published.

## Acknowledgments

This work uses data provided by patients and collected by the NHS as part of their care and support. We would also like to acknowledge all data providers who make anonymised data available for research.

We wish to acknowledge the collaborative partnership that enabled acquisition and access to the de-identified data, which led to this output. The collaboration was led by the Swansea University Health Data Research UK team under the direction of the Welsh Government Technical Advisory Cell (TAC) and includes the following groups and organisations: the Secure Anonymised Information Linkage (SAIL) Databank, Administrative Data Research (ADR) Wales, NHS Wales Informatics Service (NWIS), Public Health Wales, NHS Shared Services Partnership and the Welsh Ambulance Service Trust (WAST). All research conducted has been completed under the permission and approval of the SAIL independent Information Governance Review Panel (IGRP) project number 0911.

