## Supplementary Material for "Intensity of COVID-19 in care homes following Hospital Discharge in the early stages of the UK epidemic"

**MCMC implementation**: The minimization was performed with a Markov Chain Monte Carlo (MCMC) model implemented using PyMC3 [1]; see [2]. Half-normal distributions with mean 1 and standard deviation 1 were used as priors on all fitted variables. The Metropolis sampler was used with 2 chains; 500 initial samples discarded for thermalization, and subsequently 2000 samples are drawn from the posterior distribution. These numbers were chosen to give a well-thermalised ensemble with sufficient independent samples to be representative of the true distribution. The shape and position of the distribution were found to be consistent between independent chains, indicating good confidence that the algorithm converged.

The log-likelihood was calculated from the intensity as:

$$L = \sum_{i,t} \ln \frac{\left(\lambda_i(t)\right)^{n_i^c(t)} e^{-\lambda_i}}{n_i^c(t)!} = \sum_{i,t} n_i^c(t) \ln \lambda_i(t) - \lambda_i(t) - \ln(n_i^c(t)!)\,.$$

Both the intensity and log-likelihood calculation were implemented using Numpy (Harris et al., 2020, Bennet, 2020).

**Figure S1**. Illustration of Hawkes process model. Each care home starts with a baseline case probability (per day), in this case 0.01 per day (representing import risk in a large care home). Following a case event in the home, the self-excitation process increases the probability of further cases, representing the process of spread within the home. If no further cases occur, the overall intensity declines. The shape of the decline is determined by the distribution of the serial interval for COVID-19. If the case sparks an outbreak (“daughter events”), then they also contribute to the self-exciting process, and the intensity increases further. The effect of 3 introduction is on the same day is shown (orange), alongside the example of a cluster over several days, which tends to lengthen the risk period for an outbreak.

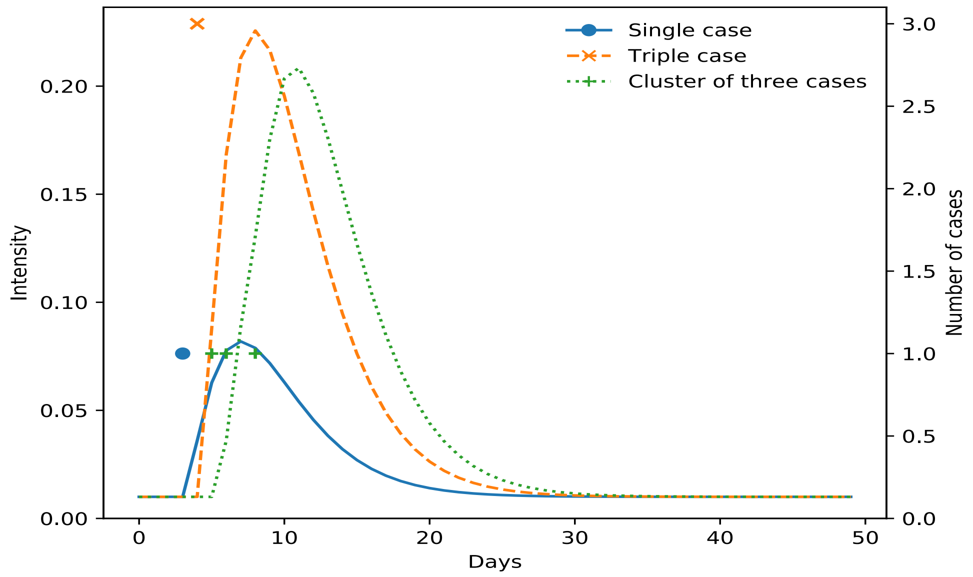

**Table S1: Summary of care home characteristics**

The care home characteristics show differences between all care homes in the Care Inspectorate Wales register and the care homes successfully linked to the WDSD. Overall, the characteristics of the linked data remained representative of the entire Care Inspectorate Wales register.

| All care homes | Number of care homes | % of total care homes | Restricted to care homes with at least one resident | Number of care homes | % of total care homes | % Linked |
|---|---|---|---|---|---|---|
| Total | 1046 | | Total | 881 | | 84.2% |
| Nursing | 253 | 24.2% | Nursing | 223 | 25.3% | 88.1% |
| Learning Disability | 439 | 42.0% | Learning Disability | 360 | 40.9% | 82.0% |
| Mental Health | 507 | 48.5% | Mental Health | 433 | 49.1% | 85.4% |
| Adult Personal Care | 988 | 94.5% | Adult Personal Care | 839 | 95.2% | 84.9% |
| Child Personal Care | 20 | 1.9% | Child Personal Care | 12 | 1.4% | 60.0% |
| $m^2$/person | | | $m^2$/person | | | |
| Mean $m^2$/person | 27.4 | | Mean $m^2$/person | 26.0 | | |
| S.D. $m^2$/person | 33.2 | | S.D. $m^2$/person | 33.1 | | |
| [1,14] | 218 | 20.8% | [1,14] | 193 | 21.9% | |
| (14,19] | 232 | 22.2% | (14,18] | 174 | 19.8% | |
| (19,25] | 202 | 19.3% | (18,24] | 183 | 20.8% | |
| (25,35] | 194 | 18.5% | (24,33] | 161 | 18.3% | |
| (35,Inf] | 197 | 18.8% | (33,845] | 170 | 19.3% | |
| Missing | 3 | 0.3% | Missing | 0 | | |
| Capacity (Maximum Number of Residents) | | | Capacity (Maximum Number of Residents) | | | |
| Mean places | 23.9 | | Mean places | 24.9 | | |
| S.D. places | 21.1 | | S.D. places | 21.2 | | |
| [0,5] | 239 | 22.8% | [1,5] | 186 | 21.1% | |
| (5,13] | 181 | 17.3% | (5,16] | 180 | 20.4% | |
| (13,26] | 208 | 19.9% | (16,28] | 176 | 20.0% | |
| (26,38] | 219 | 20.9% | (28,38] | 164 | 18.6% | |
| (38,133] | 196 | 18.7% | (38,133] | 175 | 19.9% | |
| Missing | 3 | 0.3% | Missing | 0 | 0.0% | |

**Figure S2**. Association between the first wave of the Wales COVID-19 epidemic and hospital discharge events. (1) Total increases in COVID cases in care homes, (2) Total care homes with 2 or more positive cases, (3) Number of residents recorded in the Welsh Demographic Service Dataset , (4) Number of hospital discharges from care home residents.

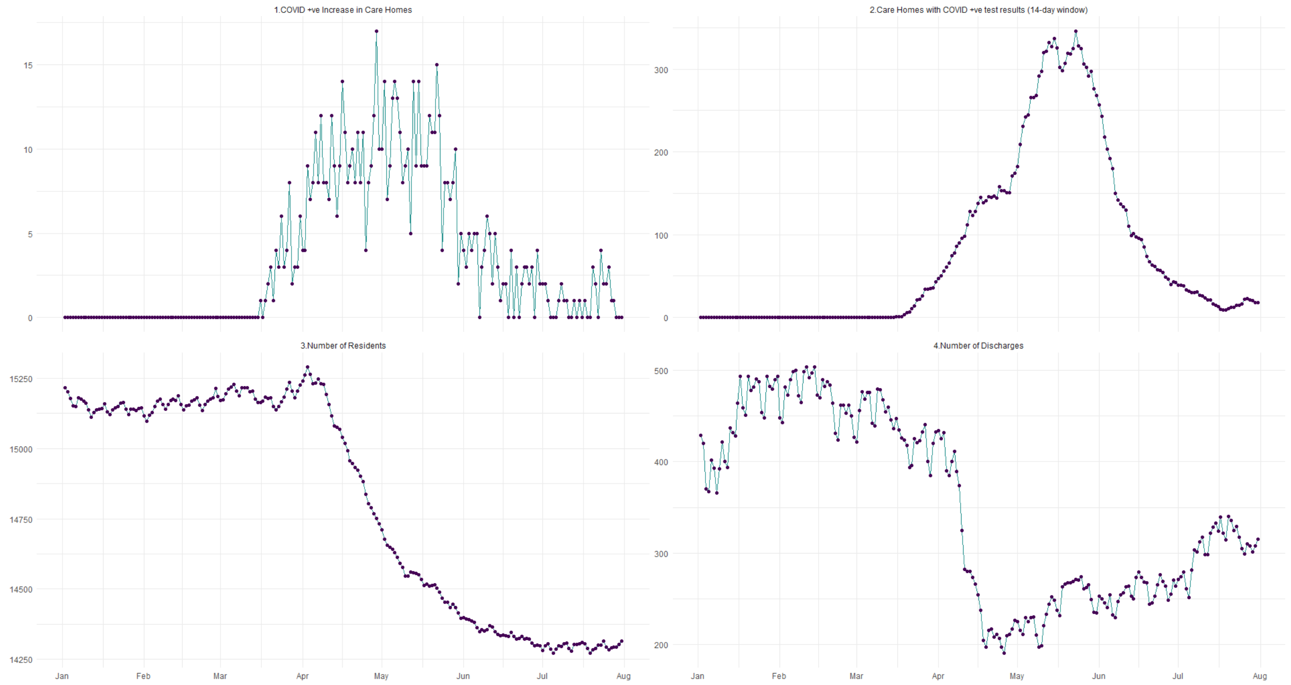

**Table S2.** Unadjusted odds ratios for the univariable multi-level logistic regression models, with the dependent variable being an increase in COVID-19 positive cases in a care home.

| **Odds Ratios** | | | | | | | |
|---|---|---|---|---|---|---|---|
| **Intercept** | 0.004 (0.003,0.005) | 0.004 (0.003,0.004) | 0.003 (0.002,0.003) | 0.006 (0.005,0.007) | 0.005 (0.004,0.006) | 0.005 (0.004,0.007) | 0 (0,0.001) |
| **Hospital Discharge** | | | | | | | |
| 1-day lagged | - | 1.237 (1.009,1.518) | - | - | - | - | - |
| **Care Home Services** | | | | | | | |
| Nursing | - | - | 3.03 (2.241,4.097) | - | - | - | - |
| Learning Disabilities | - | - | - | 0.248 (0.183,0.337) | - | - | - |
| Mental Health | - | - | - | - | 0.681 (0.515,0.899) | - | - |
| $m^2$ per person | | | | | | | |
| $m^2$(14,18] | - | - | - | - | - | 1.044 (0.698,1.562) | - |
| $m^2$(18,24] | - | - | - | - | - | 0.79 (0.527,1.185) | - |
| $m^2$(24,33] | - | - | - | - | - | 0.605 (0.393,0.932) | - |
| $m^2$(33,845] | - | - | - | - | - | 0.43 (0.276,0.669) | - |
| **Capacity** | | | | | | | |
| places(5,16] | - | - | - | - | - | - | 2.533 (1.226,5.232) |
| places(16,28] | - | - | - | - | - | - | 10.732 (5.566,20.692) |
| places(28,38] | - | - | - | - | - | - | 16.893 (8.833,32.308) |
| places(38,133] | - | - | - | - | - | - | 34.662 (18.342,65.503) |
| **Random Effects** | | | | | | | |
| Month | 9.091 (8.424,9.758) | 9.192 (8.51,9.873) | 8.151 (7.494,8.808) | 7.819 (7.182,8.456) | 9.069 (8.399,9.738) | 8.817 (8.157,9.477) | 5.894 (5.325,6.463) |
| Care Homes | 1.744 (1.332,2.156) | 1.559 (1.158,1.961) | 1.801 (1.378,2.224) | 1.608 (1.205,2.012) | 1.735 (1.321,2.148) | 1.647 (1.244,2.051) | 0.828 (0.511,1.145) |
| Local Authority | 0.152 (-0.006,0.31) | 0.145 (-0.006,0.296) | 0.172 (0,0.345) | 0.129 (-0.014,0.273) | 0.14 (-0.01,0.29) | 0.108 (-0.021,0.237) | 0.09 (-0.021,0.2) |
| **-** | - | - | - | - | - | - | - |
| **Observations** | 186772 | 186772 | 186772 | 186772 | 186772 | 186772 | 186772 |
| Months (Jan - July) | 7 | 7 | 7 | 7 | 7 | 7 | 7 |
| Care Homes | 881 | 881 | 881 | 881 | 881 | 881 | 881 |
| Local Authorities | 22 | 22 | 22 | 22 | 22 | 22 | 22 |

**Figure S3**. MCMC estimated posterior distributions, and full trace, for all parameters of full Hawkes models.

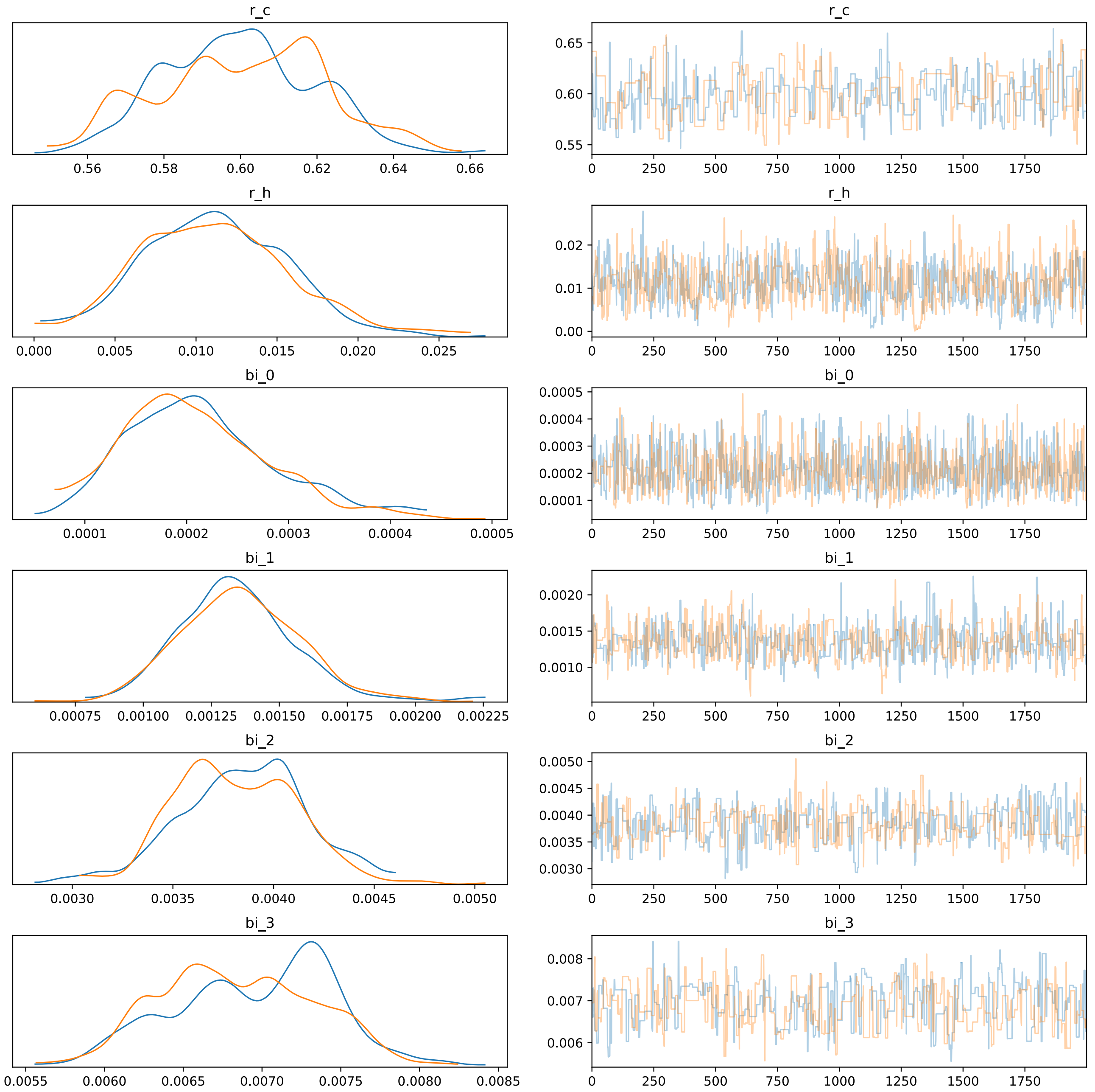

**Figure S4.** Illustration of magnitude of hospital discharge estimate. This figure shows the estimated baseline intensity for the four quartiles of care home size. Starting at these baseline levels, we simulated the Hawkes process using a hospital discharge every week. This therefore represents a very high potential exposure of the home (much higher than the average number experienced in the cohort). The effect of weekly exposure can be seen to be very similar to the difference between a Q1 and Q2 care home. The effect of weekly exposure is much less than the difference between a Q2 and Q3 care home, and between a Q3 and Q4.

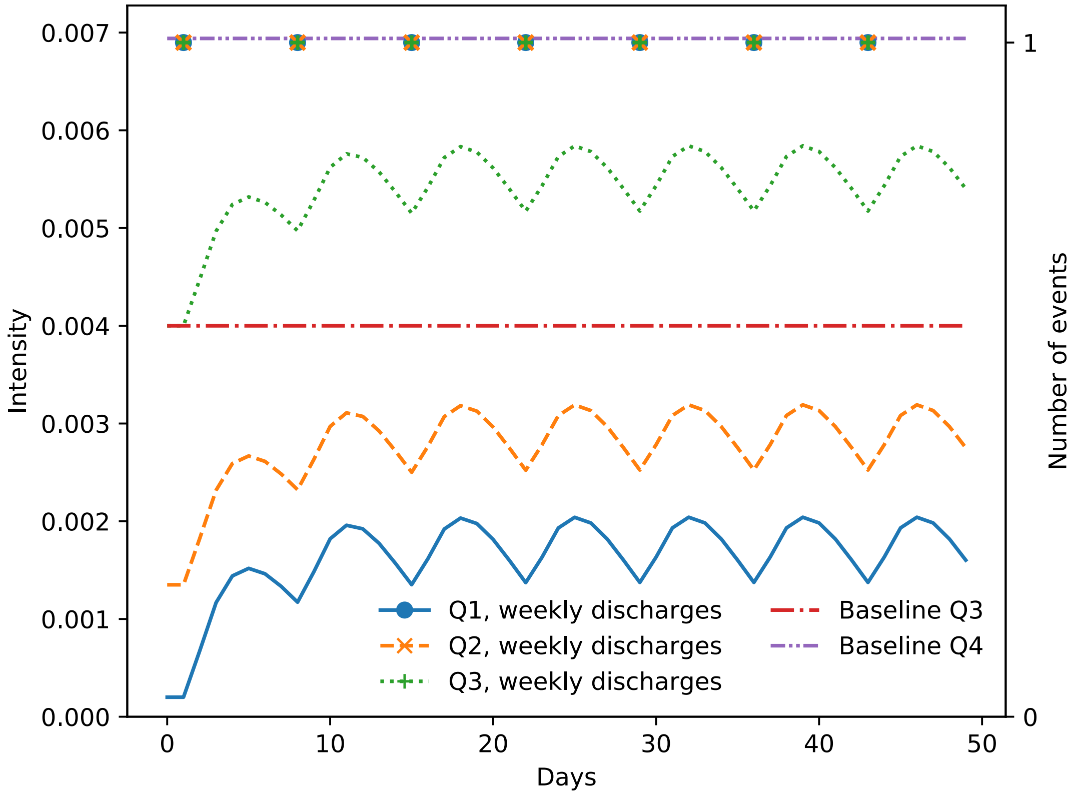


[1] J. Salvatier, T. V. Wiecki, and C. Fonnesbeck, "Probabilistic programming in Python using PyMC3," *PeerJ Comput. Sci.*, vol. 2016, no. 4, p. e55, Apr. 2016, doi: 10.7717/peerj-cs.55.

[2] E. Bennett, "GitHub - sa2c/care-home-fit: Tool to fit care home data in the SAIL environment to measure the impact of hospital transfers on infections." 2020.

[3] C. R. Harris *et al.*, "Array programming with NumPy," *Nature*, vol. 585, no. 7825, pp. 357–362, 2020, doi: 10.1038/s41586-020-2649-2.
